# Mitochondrial dysfunction in PTSD: A mechanism to understand trauma susceptibility?

**DOI:** 10.1101/2025.03.28.25324821

**Authors:** Felippe E. Amorim, Charlotte S. Rye, Amy L. Milton

## Abstract

**Rationale and Objectives:** Post-Traumatic Stress Disorder (PTSD) is a complex mental health condition that arises following exposure to traumatic events. Converging evidence suggests mitochondrial dysfunction and brain energy metabolism impairment in its pathophysiology. Thus, integrating mitochondrial data from both preclinical and clinical studies may help us to have a deeper understanding of the pathophysiological mechanisms underlying PTSD.

**Methods:** Using PubMed, Scopus and Web of Science online databases, we conducted a search for peer-reviewed manuscripts targeting both mitochondrial-related activity and PTSD. Our search yielded 43 studies in total, including 29 in rodent models and 15 clinical studies.

**Results:** Preclinical studies reported a decrease in energy metabolism with a reduction in adenosine triphosphate (ATP), impairment on the glycolytic pathway, citric acid cycle and oxidative phosphorylation pathways increasing oxidative stress and neuronal apoptosis in the brain, or systemically. In the clinical setting, studies identified 1108 participants with PTSD and 312 with partial PTSD, with these individuals showing alterations in energy production, mitochondrial DNA copy number (mtDNAcn) and elevated oxidative stress. Risperidone and AC-5216 - a selective modulator of the GABA-A receptor - emerged as potential treatments.

**Conclusion:** Our synthesis of the published findings indicates a notable overlap between results from both animal models and humans which could show a potential usage of mitochondrial-related targets as biomarkers or for drug discovery. Additionally, these results highlight the need for future research in describing whether mitochondrial dysfunction is a cause or a symptom of PTSD.

## INTRODUCTION

Stress-related illnesses, including post-traumatic stress disorder (PTSD), are not only a significant financial burden, with estimated healthcare costs exceeding £10 billion annually in the UK (Stewart, 2021), but they also cause profound personal suffering. PTSD is precipitated by either direct or indirect exposure to a traumatic event involving actual or threatened death, injury or violence, and is characterised by symptoms such as flashbacks, nightmares, intrusive memories, emotional numbness, hypervigilance, and avoidance (American Psychiatric Association, 2013). These emotional and psychological symptoms can severely impact a person’s daily functioning, affecting their relationships, work, and overall well-being. As a result, there has been a growing effort in recent decades to better understand the complex mechanisms of PTSD, aiming to alleviate both the economic and personal impacts of the disorder.

In addition to these severe and debilitating psychological symptoms, individuals with PTSD also experience high rates of physical comorbidity, with increased rates of cardiovascular disease, arthritis, asthma and gastritis (Pietrzak et al., 2012). These comorbidities not only worsen overall health burden and compound the suffering of those affected but may also be associated with poorer treatment response (Ryder et al., 2018).

While the evidence linking PTSD to physical morbidity and mortality is strong, the underlying mechanisms are largely unknown. Some conditions may result directly from lifestyle changes linked to PTSD, including reduced physical activity, substance use and medication (Zen et al., 2012). However, it has also been suggested that the underlying biological mechanisms of PTSD itself might contribute to an increased risk of somatic illness (Levine et al., 2013; Lindqvist et al., 2017; Yehuda et al., 2011). If true, PTSD may be better conceptualised as a systemic (rather than psychiatric) condition (McFarlane, 2017).

### Energy metabolism and mitochondrial alterations in stress

One physiological aspect of the brain that is often overlooked is its high demand on energy. The majority of brain metabolic processes use glucose which enters cells through glucose transporters (GLUTs) as a source for energy production (Vannuci et al., 1997). Later, this carbohydrate is phosphorylated into glucose-6-phosphate, and enters different metabolic pathways such as glycolysis and the pentose phosphate pathway. Additionally, the respiratory chain in mitochondria produces the majority of reactive oxygen species (ROS), a major source of oxidative stress (Angelova and Abramov, 2018). Despite showing negative effects such as triggering apoptotic cell death (Redza-Dutordoir and Averill-Bates, 2016), ROS also seem to be important for brain functions such as neuronal plasticity and memory (Hidalgo and Arias-Cavieres, 2016). Therefore, ROS levels need to be properly balanced by the oxidative phosphorylation (OXPHOS) function, to determine whether the beneficial signaling roles of ROS outweigh their potential to induce oxidative stress.

Interestingly, due to cellular heterogeneity, each central nervous system cell has a different metabolic profile. For example, neurons are more dependent on OXPHOS, whereas astrocytes exhibit a more glycolytic phenotype, consistent with their role in supporting neuronal activity (Belanger et al., 2011; Magistretti and Allaman, 2015). A key contributor to this neuronal phenotype is the sustained proteasomal degradation of 6-phosphofructo-2-kinase/fructose-2,6-bisphosphatase 3 (Pfkfb3), a critical regulator of glycolysis (Herrero-Mendez et al., 2009). Consequently, glucose-6 seems to be predominantly metabolized via the pentose phosphate pathway, which generates NADPH, a scavenger for neutralizing reactive oxygen species arising from OXPHOS. On the other hand, astrocytes metabolize glucose mostly via glycolysis, where most produced lactate is released extracellularly (Bolanos et al., 1994). Later, this lactate originating from astrocytes is internalized and oxidized by neurons in a process defined as astrocyte-neuron lactate shuttle (Pellerin et al., 1998; Brooks 2018). Besides providing energy, this lactate transport has a role in long-term memory formation and synaptic plasticity (Calì et al., 2019)

Under acute stress, such as exercise and ‘fight or flight’ behaviour, physiological adaptations prioritize energy redistribution, ensuring the preservation of functions critical to survival. Mitochondria, as a central regulator of energy metabolism, modulate OXPHOS efficiency and activate ancillary metabolic pathways, such as fatty acid oxidation and the urea cycle, to accommodate the altered energetic demands (Manoli et al., 2007). Physical exercise has been shown to enhance mitochondrial function in the brain by upregulating the expression of TCA cycle enzymes and respiratory chain proteins, suggesting an improved bioenergetic capacity (Navarro et al., 2004; Marques-Aleixo et al., 2015). Nonetheless, neurons seem to be uniquely constrained in their metabolic flexibility with evidence indicating that they are unable to increase their glycolytic rate during intense neuronal activity (Chuquet et al., 2010). Furthermore, neurons are unable to compensate for impaired OXPHOS by increasing their glycolytic rate. While astrocytes are capable of upregulating glycolysis to prevent ATP depletion when respiration is inhibited, neurons progressively reduce ATP levels and apoptotic death occurs (Almeida et al., 2001). One key reason for this is the low level of pfkfb3 in neurons. Transfection of neurons with pfkfb3 can increase glycolysis; however, this is followed by lactate accumulation, enhanced ROS formation and induced apoptosis (Herrero-Mendez et al., 2009). Thus, one hypothesis is that glucose metabolism via pentose phosphate pathway is used as a robust antioxidant system (Bolanos et al., 2010). Nonetheless, this metabolic rigidity renders neurons particularly vulnerable to stress-induced disruptions in mitochondrial function, with potential implications for stress-related neuropathologies.

### Mitochondria and psychiatric disorders

Many investigators have debated the association between metabolic dysfunction and psychiatric disorders (Fattal 2006, Pan et al., 2012; Vancampfort et al., 2013; Bergman and Ben-Shachar, 2016), including PTSD (Lushchak et al., 2023). This disorder shares multiple biological phenotypes associated with metabolic syndromes, such as changes in the hypothalamic-pituitary-adrenal axis, sympathetic nervous system, metabolic characteristics and neuroinflammation (Michopoulos et al., 2016). Indeed, one meta-analysis found that PTSD patients have a higher prevalence of hyperglycemia, obesity, and almost double the relative risk of having a metabolic syndrome than the general population (Rosenbaum et al., 2015). At the brain level, positron emission tomography (PET) scans of PTSD patients show different glucose absorption characteristics in several structures including the prefrontal cortex (PFC), insula and hippocampus (Molina et al., 2010), suggesting an association between disruption of glucose metabolism and the disorder.

When investigating potential metabolic mechanisms impaired in PTSD, mitochondria emerge as a primary candidate, given their role as ‘first responder’ to stress stimuli (Manoli et al., 2007). In preclinical models, exposure to predators elicits PTSD-like phenotypes in rats, and these are associated with elevated ROS levels in the blood, prefrontal cortex (PFC) and hippocampus of these animals (Wilson et al., 2013). Consistently, other PTSD-like models showed an increase in apoptosis in the medial PFC, hippocampus and amygdala (Jia et al., 2018). Additionally, a substantial amount of genes related to mitochondrial function appear to be dysregulated in the amygdala of PTSD animal models (Zhang et al., 2015).

Investigations in PTSD patients also provide corroborating evidence of mitochondrial dysfunction. Metabolomic profiling of male combat veterans with PTSD reveals an increased glycolytic rate, suggesting changes in the TCA cycle with elevated levels of lactate and pyruvate and a decrease in citrate (Mellon et al., 2019). These findings suggest an upregulation of anaerobic, and downregulation of aerobic, respiration in patients. Additionally, gene expression analyses of post-mortem PTSD brains identify a substantial amount of dysregulated genes associated with mitochondrial dysfunction, OXPHOS, citrate cycle and apoptosis (Su et al., 2008).

### Lipid Peroxidation

Lipid peroxidation is a well-known source of reactive oxygen species (ROS) (Bilici et al., 2001) and has received a great deal of attention in connection with oxidative stress (Niki et al., 2005). The process involves the breakdown of polyunsaturated fatty acids (PUFAs) within cell membranes by ROS such as hydroxyl radicals (•OH), superoxide anion (O₂•⁻), or hydrogen peroxide (H₂O₂), leading to the formation of lipid hydroperoxides (LPOs) (Augusto et al., 2002). This oxidative degradation of lipids is a key aspect of oxidative stress, which occurs when the production of ROS exceeds the body’s antioxidant defences (Preiser, 2012).

Mitochondria are particularly vulnerable to lipid peroxidation because the inner mitochondrial membrane is rich in polyunsaturated lipids (Horton et al., 1987). Lipid peroxidation in this membrane can disrupt membrane integrity, leading to a loss of membrane potential and impaired function of the respiratory chain complexes. This can cause a reduction in ATP production, resulting in cellular energy deficits (Berson et al., 1998). In addition, mitochondria are the primary site of ROS production (Chen et al., 2003), and their dysfunction can result in elevated ROS generation which further damages mitochondrial structures, setting up a vicious cycle of oxidative stress. This cycle is suggested to be particularly harmful in organs such as the brain where cell turnover is low or absent and high quantities of lipid peroxidation products can accumulate to exacerbate PTSD symptoms (Negre-Salvayre et al., 2010).

### Mitochondrial Gene Expression

Mitochondrial DNA encodes 37 genes essential to energy production (Montier et al., 2009) and mtDNAcn is matched to the energy needs of the tissue/organ (Bentlage & Attardi, 1996; Matilainen et al., 2017). Measuring mtDNAcn therefore provides a way of indirectly measuring mitochondrial functioning and variations in cellular bioenergetics, and altered mtDNAcn has been associated with a vast array of somatic disease (Johannsen & Ravussin, 2009).

Taken together, evidence from both animal and human studies highlights mitochondria as a potential target to be dysregulated in PTSD. Given the implications for understanding the pathophysiology and developing potential treatments for PTSD, this systematic review analysed the results referenced on several databases to further investigate mitochondrial processes implicated in the disorder symptomatology.

## METHODS

The protocol related to this literature review was registered prior to data collection on the Open Science Framework (OSF) (https://doi.org/10.17605/OSF.IO/9AZ8P). This review adhered to the Preferred Reporting Items for Systematic Reviews and Meta-Analyses (PRISMA) checklist (**Figure 1**; Page et al., 2021).

**Fig. 1.**
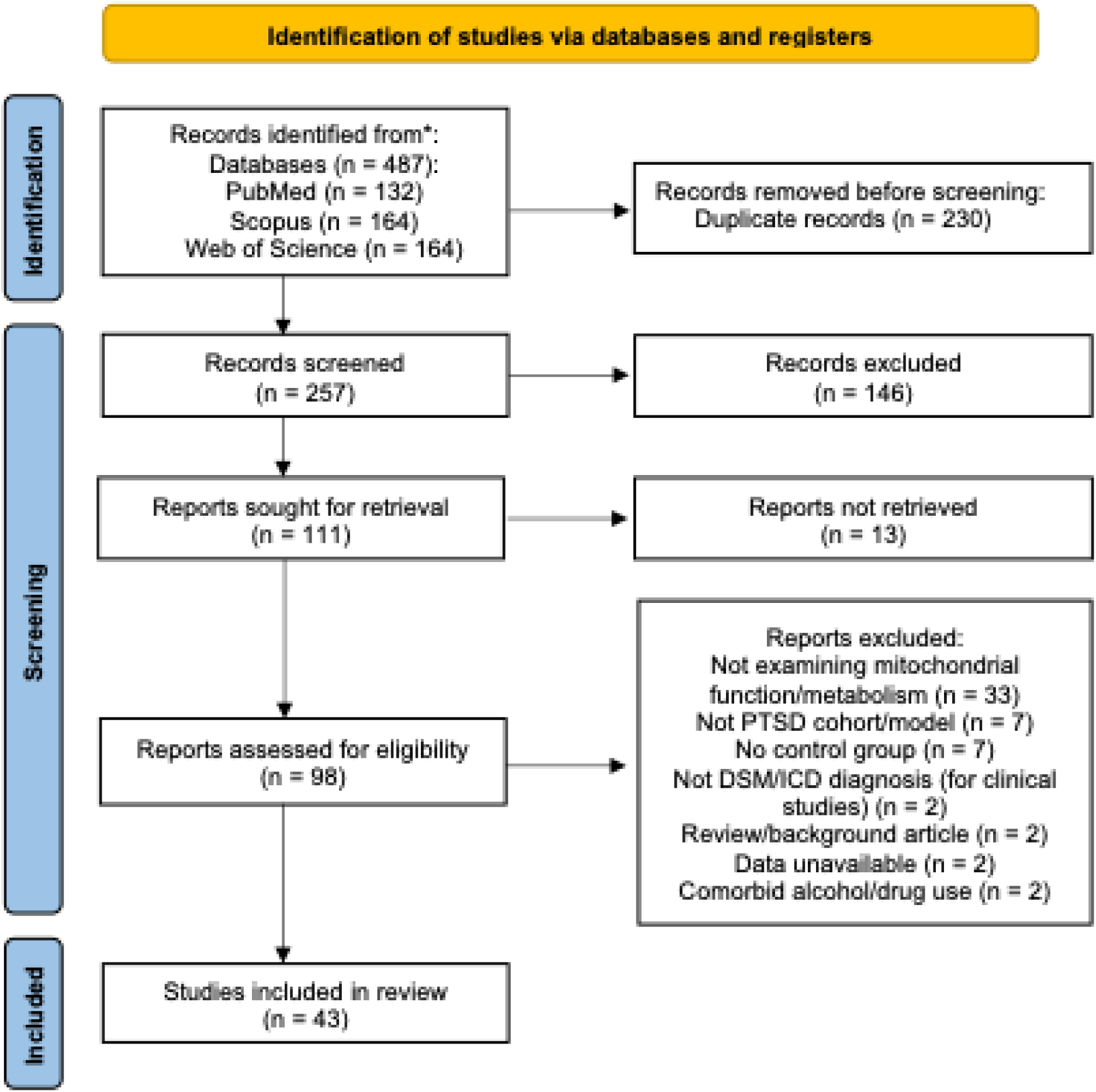
PRISMA flowchart of the literature search and study selection process. From 487 identified records, 43 primary studies of mitochondrial functioning in PTSD met the criteria for inclusion in the literature review.

### Literature Search

Thorough searches of PubMed, Scopus and Web of Science (-October 2024) were conducted, and the reference lists of any included studies were screened to identify any additional relevant studies. The following keywords were used for the search: ((mitochondri* OR Energy metabolism OR Pyruvate dehydrogenase OR respiratory chain OR Oxidative phosphorylation OR ATP OR ADP OR TCA cycle OR citric acid cycle) AND (PTSD OR post-traumatic stress)). For the initial screening, two independent investigators (C.S.R. and F.E.A.) screened studies using information available in titles and abstracts.

### Study Inclusion Criteria

Inclusion criteria were determined separately for preclinical and clinical studies. For preclinical studies, the following inclusion criteria had to be met: (i) articles employing a behavioural mouse or rat model of PTSD (ii) articles with a comparison group (either trauma-exposed, or non-exposed controls); (iii) primary studies reported in an English language peer-reviewed journal.

For clinical studies to be included, the following criteria had to be met: (i) articles on patients diagnosed with PTSD according to the Diagnostic and Statistical Manual of Mental Disorders, Fifth Edition (DSM-V) (American Psychiatric Association, 2013) or Fourth Edition (DSM-IV/DSM-IVR) (American Psychiatric Association, 1994) criteria who had no co-occurring alcohol or drug misuse; (ii) articles with a comparison group (either non-PTSD trauma-exposed, or non-exposed controls); (iii) primary studies reported in an English language peer-reviewed journal. Articles were excluded from the analysis if: (i) the publication included data that overlapped with another; (ii) there was no comparison group; (iii) data was unavailable even after contacting the corresponding author. From an initially identified 487 English language, peer-reviewed articles, 43 primary studies, containing 29 pre-clinical and 15 clinical datasets, were included in the analysis (**Figure 1**). All included studies were approved by ethical committees at the respective institutions, and protocols complied with national legislation.

## RESULTS AND DISCUSSION

### Preclinical Models

The 29 selected preclinical studies could be separated into two different categories: one investigating brain tissues such as the anterior cingulate cortex (ACC), amygdala, cerebellum, cortex, dorsal raphe nucleus, hippocampus, hypothalamus, nucleus accumbens (NAc), prefrontal cortex, prelimbic cortex (PrL) and thalamus (**Table 1**), and a one taking a systemic approach, investigating both plasma and tissues such as liver and skeletal muscle. In all studies, no female rodents were used.

**Table 1:**
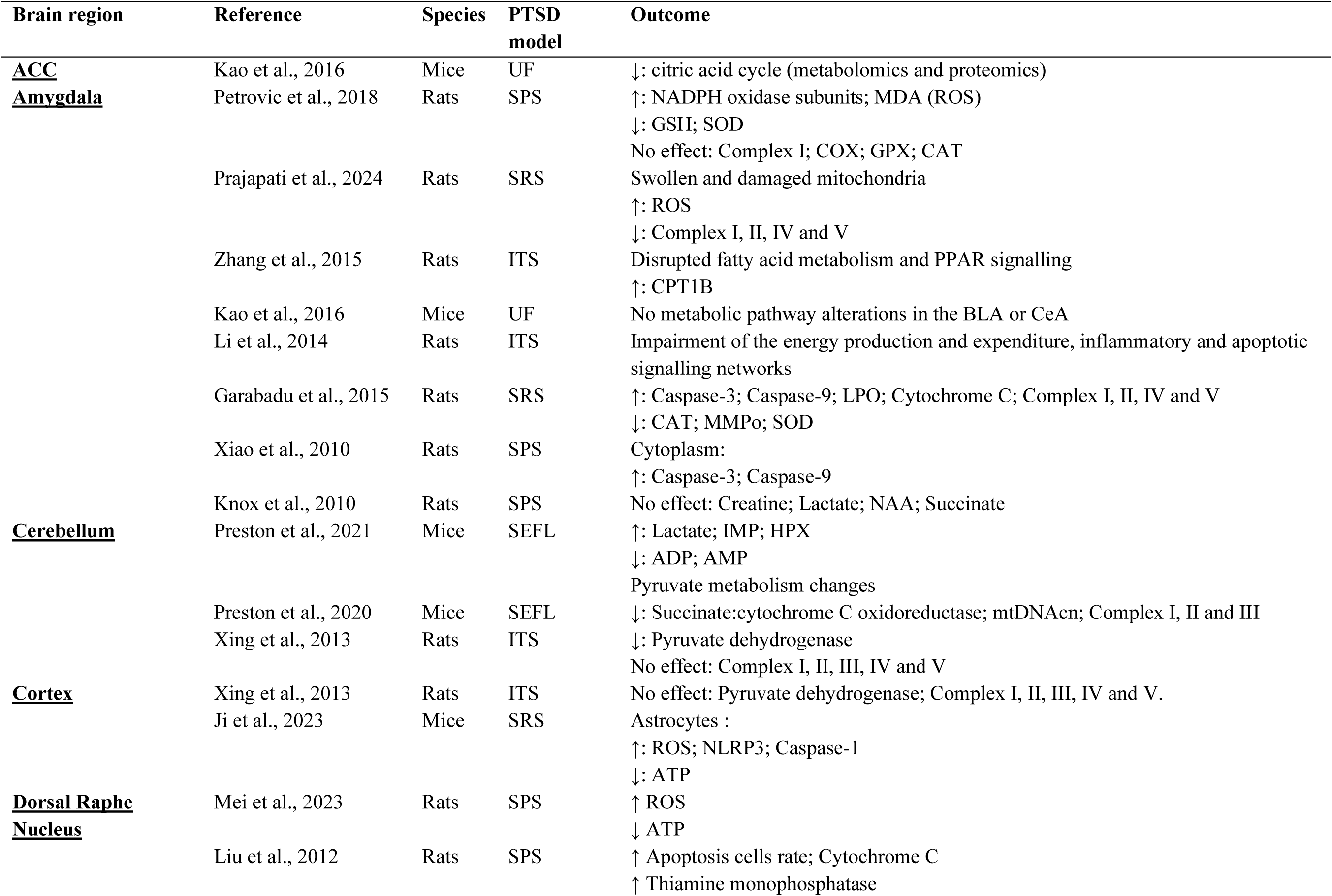
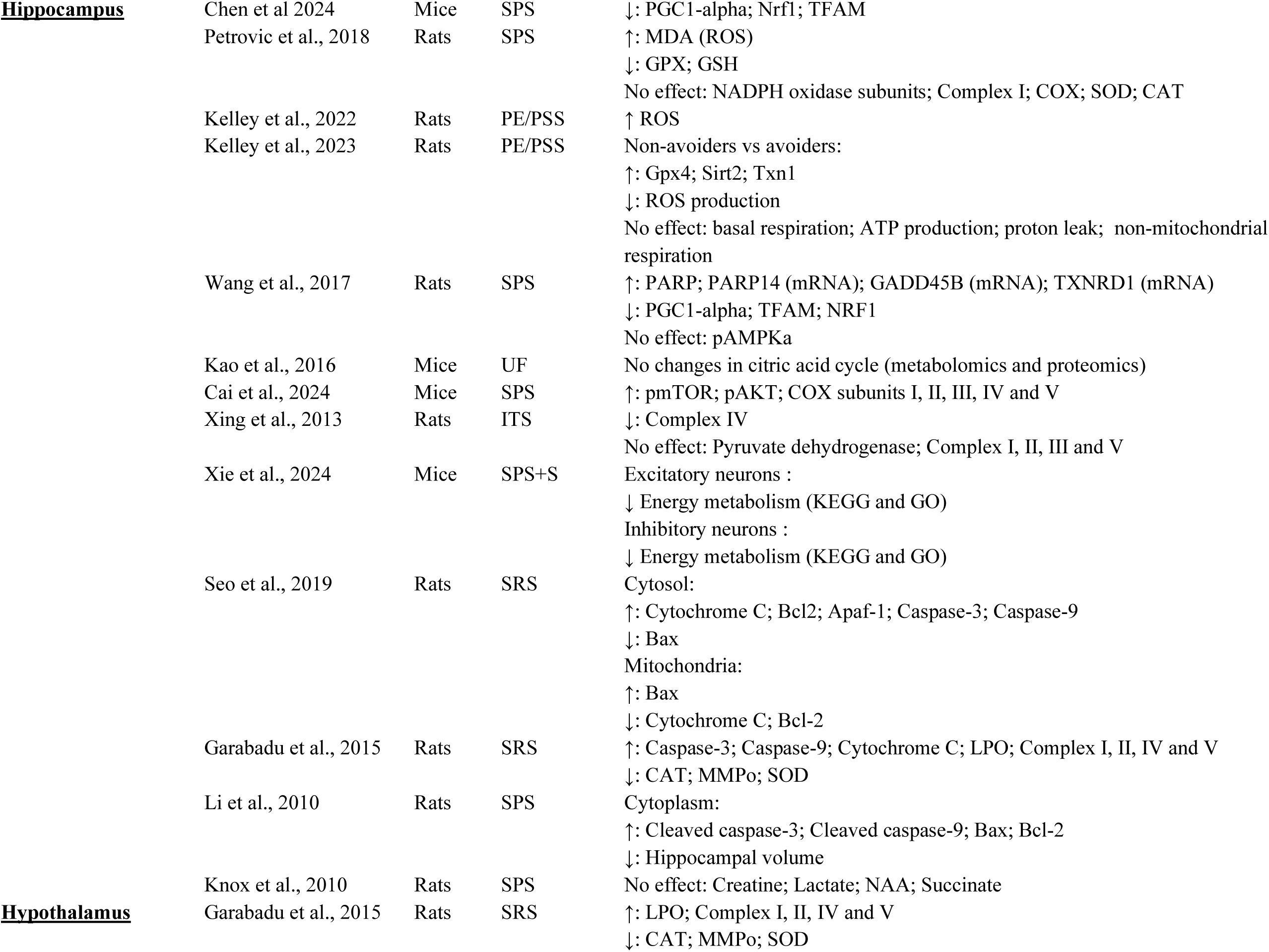
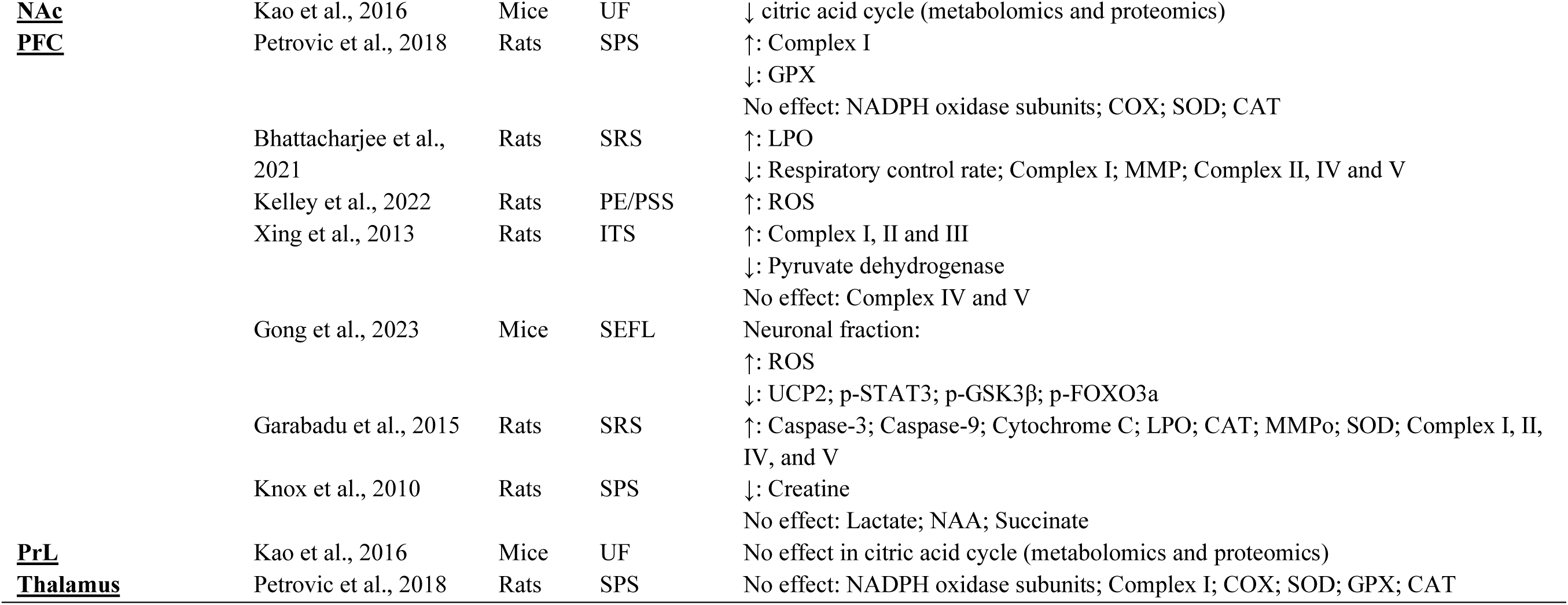
Preclinical Studies. Abbreviations: (ACC) anterior cingulate cortex, (ADP): adenosine diphosphate, (AMP) adenosine monophosphate, (Apaf-1) apoptotic protease activating factor 1, (BLA) basolateral amygdala, (CAT) catalase, (CeA) Central amygdala, (COX) cytochrome C oxidase, (GPX) glutathione peroxidase, (GSH) Glutathione, (HPX) hypoxanthine, (IMP) inosine monophosphate, (ITS) inescapable tail shock, (LPO) lipid peroxidation, (MMP) mitochondrial membrane protein, (MMPo) mitochondrial membrane potential, (NAA) N-acetylaspartate, (NAc) nucleus accumbens, (NLRP3) nucleotide-binding domain and leucine-rich repeat protein-3, (Nrf1) nuclear respiratory factor-1, (pAMPKa) phosphorylation of AMPK, (PE/PSS) predator exposure + psychosocial stress, (PFC) prefrontal cortex, (PGC1) peroxisome proliferator-activated receptor gamma coactivator-1, (pmTOR) phospho-mammalian target of rapamycin, (PPAR) peroxisome proliferator-activated receptors, (PrL) prelimbic cortex, (SOD) superoxide dismutase, (SPS) single prolonged stress, (SPS+S) single prolonged stress plus shock, (SRS) stress-re-stress, (TFAM) mitochondrial transcription factor A, and (UF) unsignalled footshock.

### Brain tissue

#### Glucose metabolism

Three studies focused on ATP levels in the brain (Ji et al., 2023; Kelley et al., 2023; Mei et al., 2023). In the first, seven days after a single-prolonged stress (SPS) protocol, rats had reduced ATP levels in the dorsal raphe nucleus compared to controls (Mei et al., 2023). Another PTSD-like mouse model showed reduced ATP levels in astrocytes throughout the cerebral cortex (Ji et al., 2023). Furthermore, Kelley and colleagues (2023) compared gene expression between non-stressed control, non-avoiders and avoiders using a predator exposure/psychosocial stress rat model. In this paradigm, avoiders exhibit a PTSD-like phenotype, whereas non-avoiders are resilient.. Interestingly, avoiders showed an upregulation in ATP synthesis and basal respiration when compared to controls and non-avoiders. However, when the oxygen consumption rate during mitochondrial respiration was measured, no statistical difference in ATP production and basal respiration was found between these groups.

Glycolysis in aerobic or anaerobic conditions uses pyruvate or lactate for ATP production which is then used to produce energy. Additionally, creatine/phosphocreatine and 5′-adenosine monophosphate-activated protein kinase (AMPK) play key roles in energy metabolism and transduction. Seven studies evaluated the energy production metabolites in different brain regions (Knox et al., 2010; Xing et al., 2013; Li et al., 2014; Kao et al., 2016; Wang et al., 2017; Preston et al., 2021; Xie et al., 2024). Xie and colleagues (2024) reported a decrease in energy metabolism within excitatory and inhibitory neurons of the hippocampus in mice following single prolonged stress plus shock (SPS+S) through gene ontology (GO) enrichment, KEGG pathway and gene set enrichment (GSE) analyses. Similarly, microarray analysis in a rat model of inescapable tail shock (ITS) evaluated 14 days after stress indicated impairment of energy production and expenditure in the amygdala (Li et al., 2014). Genes associated with ATP production, such as Idh3a and ATP5C1, were upregulated, while Glud1 - important for the citric acid cycle - was downregulated. Thus, it seems that energy production pathways are impaired within several different brain regions related to threat learning, leading to maladaptive threat expression (Alexandra Kredlow, et al., 2022).

Pyruvate dehydrogenase (PDH) serves as a mediator between anaerobic and aerobic metabolism by converting pyruvate into acetyl CoA to enter the citric acid cycle. A reduction in its activity leads to a shift to anaerobic metabolism, increasing lactate production. One study investigated PDH in four different brain regions in a rat ITS model (Xing et al., 2013). Downregulation of the enzyme was observed in the cerebellum and PFC, but no differences were found in a homogenate cerebral cortex or hippocampus compared to non-shock controls. Kao and colleagues (2016) also reported downregulation of metabolic pathways related to PDH complex in the nucleus accumbens and anterior cingulate cortex. Moreover, an integrative metabolomic analysis identified alterations in pyruvate metabolism within the cerebellum in a mouse model of stress-enhanced fear learning (Preston et al., 2021). The same study also found increased cerebellar lactate levels using nuclear magnetic resonance spectroscopy. However, in an SPS rat model, no metabolic changes were observed in the amygdala, hippocampus or PFC (Knox et al., 2010). These results are consistent with previous studies of PDH activity deficiency. For instance, mutant mice with brain-specific PDH activity showed reduced glucose oxidation, neuronal activity and an impairment of neuronal inhibition (Jakkamsetti et al., 2019). Furthermore, some studies suggest that a decline in its activity leads to a decline in cognitive functions observed in neurodegenerative disorders such as Alzheimer’s disease (Bubber et al., 2005) and Huntington’s disease (Dubinsky et al., 2017). This effect is also hypothesized to reduce glucose and oxygen consumption after brain ischemia (Martin et al., 2005).

Furthermore, Knox et al (2010) reported decreased creatine levels in the PFC, though no changes were observed in the amygdala or hippocampus. The functional implications of region-specific creatine depletion in the brain remain unclear. However, a creatine transporter deficiency mouse model, characterised by reduced cerebral creatine levels, exhibits decreased body weight and cognitive deficits in spatial, object recognition, contextual, and emotional memory tasks (Baroncelli et al., 2014; Skelton et al., 2011).

Lastly, one study reported no changes in the hippocampal phosphorylated AMPK of SPS rats measured with immunoreactivity (Wang et al., 2017). However, while mRNA expression of peroxisome proliferator-activated receptor gamma coactivator 1-alpha (PGC1a), a transcriptional activator regulated by AMPK, was downregulated in the hippocampus 1 day after stress, no changes were observed in protein expression measured with western blotting. Evidence from chronic stress rodent models suggests that hippocampal AMPK plays a key role in cognitive functions, with its reduction being associated with impairments in spatial learning and memory (Cao et al., 2014; Kim et al., 2016).

#### Impairment on the respiratory chain and mitochondrial oxidative stress in the brain

Impairment of brain mitochondria function in PTSD derives mainly from exposure to prolonged stress, either in a single or multiple sessions. Seven studies have reported some form of mitochondrial dysfunction or disruption in mitochondrial dynamics in these models (Xing et al., 2013; Garabadu et al., 2015; Petrovic et al., 2018; Preston et al., 2020; Bhattacharjee et al., 2021, Prajapati et al., 2024, Xie et al., 2024).

One of the most studied pathways is OXPHOS, with investigations into all five complexes of the respiratory chain. Mixed results appeared for the five complexes of OXPHOS, with three studies reporting an increase in at least one OXPHOS complex in the amygdala, hippocampus, hypothalamus and PFC (Xing et al., 2013; Garabadu et al., 2015; Petrovic et al., 2018). In contrast, three other studies found a downregulation of these complexes in amygdala, cerebellum, hippocampus and PFC (Preston et al., 2020; Bhattacharjee et al., 2021; Prajapati et al., 2024). Additionally, Xie and colleagues (2024), as previously described, classified hippocampal neurons into excitatory or inhibitory neurons and used GO enrichment, KEGG pathway and GSE analyses. The study reported an association between both neuron phenotypes of a SPS+S mouse model with downregulated genes in ribosomal, OXPHOS, ROS and thermogenesis pathways. Thus, there is currently no consensus on the direction of an up- or downregulation of the respiratory chain complexes, though differences from controls are consistently found. The use of differing methodologies in different brain regions make direct comparisons challenging. Interestingly, two studies from the same research group using an identical PTSD-like rodent model yielded apparently contradictory results. Prajapati et al (2024) and Garabadu et al (2015) both used a stress re-stress (SRS) PTSD-like model and measured mitochondrial complexes I, II, IV and V in the amygdala using similar measurement methods. However, while the first study reported a downregulation in the activity of these complexes, the later study found them upregulated, suggesting a stress-dependent dynamic regulation of the respiratory chain complexes to balance ATP/ADP ratios and ROS production.

An impairment on the mitochondrial respiratory chain complexes can modify the redox balance, changing the production of free radicals. Ten included studies emphasized the increase on mitochondrial oxidative stress (Xing et al., 2013; Garabadu et al., 2015; Petrovic et al., 2018; Bhattacharjee et al., 2021; Kelley et al., 2022; Gong et al., 2023; Ji et al., 2023; Kelley et al., 2023; Mei et al., 2023; Prajapati et al., 2024). An increase in ROS and products of lipid peroxidation were observed in amygdala, cortex, dorsal raphe nucleus, hippocampus, hypothalamus and PFC (Garabadu et al., 2015; Petrovic et al., 2018; Bhattacharjee et al., 2021; Kelley et al., 2022; Gong et al., 2023; Ji et al., 2023; Mei et al., 2023; Prajapati et al., 2024). Simultaneously, antioxidants such as catalase (CAT), Glutathione (GSH), glutathione peroxidase (GPx), and superoxide dismutase (SOD) and their regulators were also reduced in several brain structures such as amygdala, hippocampus, hypothalamus and PFC (Garabadu et al., 2015; Petrovic et al., 2018, Gong et al., 2023). Gong et al (2023) also observed a decrease in FoxO3a, leading to downregulation of UCP2, which regulates not only ROS production but also mitochondrial ATP production. This indicates a multiregional enhanced oxidative stress leading to an impairment to the redox balance. Additionally, only one study examined the NADPH oxidase system, finding an increase in activity in the amygdala, but no change in the hippocampus, PFC or thalamus (Petrovic et al., 2018). Thus, NOX2 does not appear to be a potential source of oxidative stress in this context. Furthermore, some studies also showed changes in only one respiratory chain complex (Xing et al., 2013; Petrovic et al., 2018). Two hypotheses emerge from these findings. The first posits that oxidative stress may not originate from respiratory chain impairment due to the complexity of mitochondrial ROS metabolism. The second suggests that up- or downregulation of a specific respiratory chain complex will lead to the development of a PTSD phenotype, as genetic variations in complex I and V have already been identified in humans (Flaquer et al., 2015).

The impairment of the redox balance, due to an increase in ROS, can potentially trigger apoptosis. Studies of PTSD using animal models have also shown an increase in neuroinflammation. All studies that evaluated neuroinflammation due to apoptosis showed an upregulation of biomarkers such as caspase-1, -3 and -9 (Li et al., 2010; Xiao et al., 2010; Garabadu et al., 2015; Seo et al., 2019, Gong et al., 2023). These three caspases are critical mediators of the inflammatory process where an upregulation can lead to neuronal cell death (Denes et al., 2012). Li et al. (2014) also showed an impairment in apoptotic signalling network function in rats using mRNA expression profiling. Finally, Wang et al. (2017) reported an increase in poly(ADP-ribose) polymerase (PARP) protein levels, an apoptosis marker, along with elevated mRNA expression of three oxidative stress-related genes: PARP14, GADD45B, and TXNRD1. PARP plays a key role in microglial activation and the release of proinflammatory cytokines, with its overactivation being implicated in several neurodegenerative disorders, including Alzheimer’s disease, Huntington’s disease, Parkinson’s disease, and multiple sclerosis (Arruri et al., 2021). The neuroinflammation literature in PTSD-like models also supports an activation of apoptosis due to an upregulation of apoptosis-related factors (Dmytriv et al., 2023; Jia et al., 2018). Together, these findings suggest that PTSD-like stress in rodent models may play a critical role in inducing neuronal apoptosis.

### Systemic

Three studies analysed the body composition of PTSD rodent models. It was shown that body weight (Dille et al., 2022) together with lean mass (Jelenik et al., 2018; Cai et al., 2024) and fat mass (Dille et al., 2022) of these animals were acutely reduced. However, 3 months following chronic variable stress (CVS), these animals showed no differences in body weight, fat mass and lean mass compared to control (Jelenik et al., 2018; Dille et al., 2022).

Four studies have reported conflicting findings on fasting blood glucose levels in PTSD-like models (Jelenik et al., 2018; Dille et al., 2022; Kondashevskaya et al., 2023; Cai et al., 2024). Using the same CVS PTSD-like model, two studies showed higher fasted blood glucose levels in the liver (Jelenik et al., 2018; Dille et al., 2022) and skeletal muscle (Jelenik et al., 2018). Similarly, serum glucose was increased in a single prolonged social stress (SPSS) mouse model (Cai et al., 2024). By contrast, Kondashevskaya et al. (2023) reported hypoglycaemia in plasma samples from high-anxiety rats subjected to a predator stress model, compared with both low-anxiety and control groups. The high-anxiety phenotype also exhibited reduced liver glycogen levels. Furthermore, Dille and colleagues (2022) reported that gluconeogenesis was initially elevated in hepatocytes in the CVS mouse model, but it was lowered after 3 months. Finally, Prajapati et al. (2024) reported increased levels of orexin, a neuropeptide that directs glucose metabolism towards the citric acid cycle and OXPHOS, in both plasma and cerebrospinal fluid of a SRS rat model compared with controls.

Cai and colleagues (2024) reported an increase in serum ATP levels in the SPSS models. However, another study showed no difference in ATP production in the CVS mouse model compared to control, acutely or 3 months after the protocol (Diller et al, 2022). A comparison between controls and rats that had undergone SEFL showed a reduction in complex III and IV, and complex activity in skeletal muscle of rats (Preston et al., 2020). However, a subsequent comparison within rats that had undergone the massed shock experience, but showed resilient vs PTSD-like behaviour, showed no differences in the muscle mitochondrial respiratory chain complex activity, suggesting that the observed changes were related to the trauma experience rather than susceptibility to the PTSD-like phenotype. Furthermore, the same research group reported a metabolomic analysis of a SEFL model stressed mice which showed an upregulation of plasma citric acid cycle metabolites in comparison with control (Preston et al., 2021). Similarly to the previous study, a comparison between resilient and susceptible trauma-exposed animals showed no significant changes. Thus, on a systemic level, stress seems to promote metabolic shifts but not seems to be relevant for different PTSD phenotypes.

The accumulation of reactive oxygen species (ROS) activates pathways that induce lipid peroxidation, ultimately leading to apoptosis. Consequently, lipid peroxidation products serve as biomarkers of oxidative stress. Two studies have reported that fatty acid oxidation is also dysregulated in preclinical PTSD models (Zhang et al., 2015; Kondashevskaya et al., 2023). One study observed an overexpression in carnitine palmitoyltransferase 1B (CPT1B), an enzyme involved in fatty acid metabolism, in both the amygdala and blood plasma of ITS rat models (Zhang et al 2015). CPT1B facilitates the transport of fatty acids into mitochondria, and is also related to the peroxisome proliferator-activated receptor pathway. Additionally, Kondashevskaya and colleagues (2023) reported an increase in lipid peroxidation products in the liver of predator stress rat model with a high-anxiety phenotype. A reduction in SOD activity was also observed in both high- and low-anxiety phenotypes.

In summary, preclinical PTSD models have several different impairments on a mitochondrial level, which leads to changes in neuroinflammation, lipid peroxidation, glucose metabolism and oxidative stress. The majority of the studies showed a consensus regarding the association of PTSD with oxidative stress, leading later to apoptosis. However, contradictory results between studies show that the mechanisms behind it are still unknown.

### Clinical Studies

The identified clinical studies included 1108 individuals with full PTSD, 312 with partial PTSD (met criterion A and experienced significant symptoms, but did not meet full diagnostic criteria), 1252 trauma-exposed individuals without PTSD and 1285 healthy control subjects. Most subjects were males (82.3% PTSD; 37.18% partial PTSD; 86.66% trauma exposed controls; 63.58% healthy controls), with the most common trauma being combat exposure. A summary of the included studies is shown in **Table 2**.

**Table 2:** Demographic and clinical characteristics of the included clinical studies. *Abbreviations:* TE = trauma exposed, no PTSD; HCs = healthy controls; w/outEDS = without excessive daytime sleepiness; wEDs = with excessive daytime sleepiness; D = discovery cohort; V = validation cohort; MDD = major depressive disorder; TBI = traumatic brain injury; NA = not available.

| Author | Year | HC or TE | n Controls | n PTSD | Mean ± SD Age (years)<br>Controls | Mean ± SD Age (years) PTSD | Sex | Nature of trauma | Comorbidities | Medication |
| --- | --- | --- | --- | --- | --- | --- | --- | --- | --- | --- |
| Blalock et al. | 2024 | TE | 121 | 111 | 33.08±8.61 | 33.84±8.54 | Male | Combat | Yes (diabetes) | Yes |
| Mellon et al. | 2019 | TE | D: 51<br>V: 31 | D: 52<br>V: 31 | D: 33.69±9.03<br>V: 30.61±5.66 | D: 34.02±8.69<br>V: 31.23±5.45 | Male | Combat | Yes (MDD +<br>metabolic<br>conditions) | Yes |
| Konjevod et al. | 2021 | HC | D: 50<br>V: 52 | D: 50<br>V: 52 | D: 58.04±9.03<br>V: 63.75±7.36 | D: 58.40±8.76<br>V: 63.56±7.28 | Male | Combat | No | NA |
| Zhang et al. | 2015 | TE | 31 | 28 | 21.4±5.0 | 27.5±5.0 | Mixed | Combat | NA | NA |
| Bersani et al. | 2016 | TE | 44 | 43 | 32.16±8.55 | 34.49±8.15 | Male | Combat | Yes (MDD) | Yes |
| Dell'Osso et al. | 2010 | HC | 23 | 25 | 39.08±11.96 | 44.77±14.10± | Mixed | Mixed | NA | No |
| Hu et al. | 2023 | TE | 689 | 151 | 29.5±7.4 | 27.9±6.3 | Mixed | Combat | NA | No |
| Konjevod et al. | 2022 | HC | 241 | 235 | IQR: 42.55 – 47.00 | IQR: 51.61 – 55.00 | Male | Combat | No | No |
| Pattinson et al. | 2019 | TE | 57 | w/outEDs: 26<br>wEDs: 22 | 35.2±11.2 | w/outEDs: 37.7±9.5<br>wEDs: 43.9±10.8 | Mixed | Combat | Yes (MDD + TBI) | Yes |
| Behnke et al. | 2022 | TE | 14 | 15 | 22.5±12.7 | 24.5±8.5 | Mixed | Torture/Civil Unrest | Yes (MDD +<br>physical disease) | Yes |
| Talbot et al. | 2015 | HC | 50 | 44 | 30.34±8.11 | 30.55±6.57 | Mixed | Mixed | Yes (MDD) | NA |
| Flaquer et al. | 2015 | HC | 875 | Partial: 312<br>Full: 51 | M: 52.3<br>F: 50.8 | Partial PTSD: M: 54.2, F: 51.8<br>Full PTSD: M: 58.6, F: 52.3 | Mixed | Mixed | NA | NA |
| Su et al. | 2018 | TE | 71 | 78 | 43.4±9.9 | 43.0±10.4 | Mixed | Natural Disaster | No | No |
| Atli et al. | 2016 | Both | HC: 38<br>TE: 31 | 32 | TE: 30.6±6.1<br>HC: 32.9±11.8 | 33.1±12.4 | Mixed | Natural Disaster | Yes | NA |
| Nikolac Perković et al. | 2021 | HC | 58 | 62 | 57(median) | 56(median) | Male | Combat | No | No |

The search identified five main areas of investigation: i) altered metabolite concentrations; ii) lipid peroxidation; iii) mitochondrial DNA copy number (mtDNAcn); iv) mitochondrial gene expression; v) circulating cell-free mitochondrial DNA (ccf-mtDNA).

### Altered Metabolite Concentrations

Metabolic signatures have previously been identified for several mental health and neurodegenerative disorders (e.g., Koenig et al., 2018; Rozen et al., 2005), and it has recently been suggested that metabolic differences may also characterise PTSD (Karabatsiakis et al., 2015). Supporting this, we identified three studies reporting alterations in pathways involved in fatty acid uptake and metabolism (Konjevod et al., 2021; Mellon et al., 2019; Talbot et al., 2015). In their metabolomic analysis of male combat veterans, Mellon and colleagues (2019) reported decreased abundance of fatty acids such as eicosenoate, linolenate and docosahexaenoate in PTSD subjects. Similarly, Konjevod et al. (2021) reported decreased levels of phosphatidylcholines (PCs) – a type of glycerophospholipid that play a structural role in mitochondria (Mejia & Hatch, 2016) – in PTSD. This metabolic signature was accompanied by the observation of increased triglyceride (Talbot et al., 2015) and phosphatidylethanolamine (PE) levels in PTSD (Konjevod et al., 2021). Proper maintenance of the PC:PE ratio is important for membrane integrity, liver function and in responding to oxidative stress (Calzada et al., 2016), with decreased PC levels associated with nonalcoholic steatohepatitis (Arendt et al., 2013) and cardiovascular disease (Vianello et al., 2024). Consequently, these findings are consistent with higher metabolic risk and alterations to oxidative phosphorylation in PTSD (Calzada et al., 2016).

Whilst the nature of these metabolic differences is unknown (i.e. whether they are sequelae of PTSD or pre-existing risk factors), they are all consistent with significant alterations in mitochondrial function and energy utilisation in PTSD. Importantly, none of these alterations could be explained by comorbidities including major depressive disorder, BMI, blood glucose, sleep quality or medication usage, and were also observed in young, otherwise healthy individuals (Mellon et al., 2019; Talbot et al., 2015). It therefore appears that alterations in lipid metabolism could present the possibility of early identification of those susceptible to PTSD, and early intervention.

Our search also identified abnormal concentrations of N-acetylaspartate (NAA) and creatine in the brains of drug-naïve PTSD patients (Su et al., 2018), further supplementing the notion of altered metabolism in PTSD. Su and colleagues (2018) observed increased levels of NAA in the anterior cingulate cortex (ACC) in individuals with PTSD compared to trauma-exposed controls, suggesting potential alterations in neuronal metabolism and function within this region (Moffett et al., 2007). NAA is a compound predominantly found in neurons and is often used as a marker for neuronal integrity and health (Moffett et al., 2014). It is also primarily synthesised in mitochondria (Ariyannur et al., 2008). Elevated NAA concentrations could therefore be a marker of altered mitochondrial energetics in the ACC – either enhanced neuronal activity or a compensatory response to stress-related changes. Further research is needed to better understand the implications of these findings and whether NAA could serve as a reliable neuroimaging marker in PTSD.

Su and colleagues (2018) also reported increased creatine levels in the amygdala of individuals with PTSD, pointing to potential disruptions in cerebral energy metabolism linked to trauma-related stress responses. Creatine plays a crucial role in cellular bioenergetics by maintaining the phosphocreatine shuttle, which facilitates the rapid transfer of high-energy phosphate bonds from mitochondria to other cellular compartments to maintain synaptic function and neuroplasticity (Andres et al., 2008). Elevated creatine concentrations in the amygdala may therefore reflect heightened neural activity and the increased metabolic demands associated with PTSD (Rasmusson et al., 2010). Over time, heightened creatine concentration in the amygdala could result in mitochondrial strain, potentially leading to metabolic dysregulation (Rasmusson et al., 2010). Consequently, understanding the interplay between elevated creatine levels and mitochondrial function will be critical in offering new perspectives on how altered mitochondrial dynamics contribute to the disorder’s neural and behavioural manifestations.

### Lipid Peroxidation and ROS

The breakdown products of lipid peroxides, such as malondialdehyde (MDA) and 4-hydroxy-2-nonenal (4-HNE), can directly modify proteins, lipids, and DNA within the mitochondria, and our search revealed several studies reporting heightened levels of both markers in PTSD (Atli et al., 2016; Konjevod et al., 2022; Perković et al., 2021) as well as over-expression of CPT1B, (Zhang et al., 2015). These reactive aldehydes are highly cytotoxic and can form adducts with mitochondrial proteins and enzymes, impairing their activity. This can trigger mitochondrial dysfunction, including the release of pro-apoptotic factors like cytochrome c, leading to programmed apoptosis and necrosis (Krajewski et al., 1999). Additionally, 4-HNE has been shown to alter the permeability of the blood-brain barrier (BBB) during oxidative stress (Žarković et al., 1999), thus allowing it to enter the brain, and this may further exacerbate the vicious cycle of lipid peroxidation in the brain.

MDA and 4-HNE may also indirectly compromise mitochondrial function via activation of inflammatory pathways. These reactive aldehydes can bind to receptors like the aldehyde receptor (RAGE) or trigger the NF-kB pathway to promote the release of pro-inflammatory cytokines such as IL-1β, IL-6 and TNF-a (Pickering & O’Connor, 2007; Tak & Firestein, 2001). Neuroinflammation results in the inhibition of the neuronal 2-oxoglutarate dehydrogenase complex (OGDHC) - a tricarboxylic acid (TCA) cycle enzyme – which inhibits the uptake of glutamate by mitochondria (Vaglio-Garro et al., 2024). Consequently, there is an extracellular accumulation of glutamate which stimulates the toxic glutamate pathway mediated by GluN2B-containing NMDA receptors to result in dysregulation of intracellular redox homeostasis, ferroptosis and mitochondrial dysfunction (Weidinger et al., 2023). Impairments to the TCA cycle due to loss of OGDHC also disrupt oxidative phosphorylation, resulting in an inability of mitochondria to meet their bioenergetic demands (Weidinger et al., 2023).

Given the role of lipid peroxidation in the production of ROS (Bilici et al., 2001) and the association between ROS levels and expression of the mitochondrial translocator protein (18 kDa) (TSPO) (Barichello et al., 2017), increased ROS production in PTSD may explain the finding by Dell’Osso and colleagues (2010) of decreased TSPO density in PTSD. TSPO is an outer mitochondrial membrane (OMM) protein which is necessary for cholesterol import and steroid production (Rone et al., 2009) as well as regulation of mitochondrial quality control and haem synthesis (Barichello et al., 2017). High levels of ROS (including those produced by lipid peroxidation) prevent the expression of TSPO, which decreases cholesterol transport to the inner mitochondrial membrane and dysregulates mitochondrial homeostasis (Batarseh & Papadopoulos, 2010). Decreased TSPO expression has also been strongly associated with increased generation of pro-inflammatory cytokines (Bae et al., 2014), further exacerbating the inflammatory and bioenergetic consequences of lipid peroxidation.

### mtDNAcn

Our search identified two clinical studies examining mtDNAcn in PTSD and trauma-exposed non-PTSD controls (Bersani et al., 2016; Hu et al., 2023). However, whilst Bersani et al. (2016) reported a reduction in mtDNAcn in male combat veterans with PTSD, Hu and colleagues (2023) reported significantly higher mtDNAcn in females with PTSD than either male or female controls or males with PTSD. Furthermore, they observed no difference in mtDNAcn between males with PTSD and male or female controls. However, when methodological differences between the two studies are considered, these seemingly contradictory results can be reconciled. Bersani and colleagues (2016) observed an inverted U-shaped function to their data, with alterations in mtDNAcn only seen in moderate, and not mild or severe PTSD. Since Hu et al. (2023) did not stratify by symptom severity, significant results in the male group may have been masked.

How the cellular mtDNAcn is adjusted to and maintained at a certain level is not completely understood, but an intriguing hypothesis is that an increase in the absolute mtDNAcn could be a compensatory mechanism aimed at sustaining OXPHOS activity. Consequently, the increase in mtDNAcn in female service members with PTSD may indicate attempts to maintain OXPHOS activity to meet increased bioenergetic demands. To determine this would require correlatory analysis between symptom severity in the female PTSD group and mtDNAcn.

A reduction in mtDNAcn typically occurs with aging (Mengel-From et al., 2014), and low mtDNAcn has been associated with high mortality and poor cognitive and physical health (Filograna et al., 2021) This has significant implications for the studies reviewed here since the cohort described by Bersani et al. (2016) contains individuals with hypertension, diabetes, and angina. It is therefore impossible to disentangle the effects of PTSD on mtDNAcn versus the effects of these somatic conditions.

Consequently, despite these studies revealing a potential association between mtDNAcn, mitochondrial dysfunction and PTSD symptoms, the true nature of sex-related differences (if any) remains to be discerned. The data from Hu and colleagues (2023) appears to indicate a sex-dependent effect on mtDNAcn in PTSD, but it is well established that females typically have a higher mtDNAcn than males (Ding et al., 2015) even in the absence of disease or disorder. Furthermore, even if an association between mtDNAcn and PTSD is established, it does not necessarily imply a causal connection, particularly given the cross-sectional nature of studies to date. Indeed, both studies are cross-sectional and only include measures at a single time-point in military cohorts, and prospective longitudinal studies will be required to determine whether mtDNAcn may be a potential biomarker for PTSD.

### Mitochondrial Gene Expression

Our search identified four clinical studies reporting altered expression of mitochondrial genes in PTSD (Behnke et al., 2022; Flaquer et al., 2015; Pattinson et al., 2020; Zhang et al., 2015).

In their analysis of 978 mitochondrial single nucleotide polymorphisms (SNPs), Flaquer and colleagues (2015) obtained significant associations between PTSD and no-PTSD and two mitochondrial SNPs (mtSNPs): a missense mutation (mt8414C → T) located in ATP synthase subunit 8 (MT-ATP8) and a synonymous mutation (mt12501G → C) located in the NADH dehydrogenase subunit 5 (MT-ND5). Heteroplasmy for the two variants towards a larger number of the respective minor alleles was found to increase the risk of having PTSD regardless of age or sex (Flaquer et al., 2015). Despite the mt12501G → A mutation in MT-ND5 being synonymous and thus leading to an unchanged protein, it is hypothesised that different codons may lead to variations in protein expression levels, potentially resulting in a loss of mitochondrial membrane potential (Flaquer et al., 2015). Furthermore, the PFC may be particularly vulnerable to the increase in ROS production associated with this mutation (Andreazza et al., 2010), adding to the vicious cycle of oxidative stress and exacerbating PTSD symptoms.

Elevated ROS production induces increased DNA damage, leading cells to increase their metabolic expenditure for DNA maintenance and repair (Behnke et al., 2022). Whilst this is facilitated by various proteins acting together as a co-regulated cascade (David & Williams, 1998) including Poly [ADP-ribose] polymerase 1 (PARP1) and X-ray repair cross-complementing protein 1 (XRCC1) (Behnke et al., 2022), mitochondria do not appear to contain the full range of DNA repair mechanisms that operate in the nucleus (Liu & Demple, 2010). For example, evidence suggests that mitochondria lack the nucleotide excision repair (NER) mechanism responsible for removal of UV-induced pyrimidine dimers (Reardon et al., 1999). Impairments in cellular energy provision as a direct consequence of MT-ATP8 and/or NT-ND5 dysfunction would then further dysregulate ROS production by the mitochondrial respiratory chain (Houštěk et al., 2006), irreversibly decreasing the ability of the cell to meet their metabolic demands. PTSD may then result from a pathological persistence of this cycle that occurs even after the inciting trauma is gone.

Intriguingly, our search also suggested that expression of DNA repair genes themselves may be dysregulated in PTSD, thus further fuelling this cycle (Behnke et al., 2022). Through analysis of blood samples obtained from 15 individuals with PTSD and 14 trauma-exposed controls, Behnke and colleagues (2022) observed significantly higher XRCC1 expression in PTSD patients, and a positive association between PARP1 expression and PTSD symptoms across the entire cohort. It is well-known that PARP1-regulated DNA repair critically depends on NAD+ availability (Thomas et al., 2018) and results in a shift in cellular energy metabolism from oxidative phosphorylation to glycolysis to ensure such availability and decrease oxidative stress-related damage (Murata et al., 2019). Furthermore, lower XRCC1 and PARP1 expression appears to be implicated in faster DNA repair (Behnke et al., 2022). Consequently, whilst the observed increase in XRCC1 and PARP1 expression may initially be compensatory in nature to counteract oxidative stress-related DNA damage, it has the unwanted effect of further impairing DNA repair and cellular energy provision. When coupled with potential PTSD-relevant mutations to MT-ATP8 and/or MT-ND5 and altered expression of genes involved in oxidative phosphorylation and the TCA cycle (Zhang et al., 2015), this would ultimately result in increased DNA damage but correspondingly decreased metabolic resources for DNA maintenance and repair and increased lactate levels (Lenzen, 2014).

Intriguingly however, findings from Pattison and colleagues (2020) suggest that dysregulation of genes involved in mitochondrial production and function may be associated with specific symptoms of PTSD and not necessarily the condition itself. In their recent analysis of non-treatment seeking military personnel and veterans, Pattison et al. (2020) reported that expression of transcription factor B2, mitochondrial (TFB2M) – a gene required for the transcription of mitochondrial genes (Litonin et al., 2010) – was significantly upregulated in individuals with PTSD and comorbid excessive daytime sleepiness compared to those with PTSD who did not experience this symptom. Furthermore, there were no differences between either PTSD group and controls. These findings contradict those of the other included studies, suggesting that mitochondrial dysregulation is due to the presence of comorbidities, not PTSD itself.

However, it must be noted that the control group used by Pattinson and colleagues (2020) had Clinician Administered PTSD Scale for the DSM-5 (CAPS-5) scores ranging from 17-51, indicating some level of PTSD symptoms across groups. It therefore remains possible that altered mitochondrial gene expression does categorise at least some cases of PTSD.

### Circulating Cell-Free Mitochondrial DNA Levels (ccf-mtDNA)

Ccf-mtDNA is derived and released from cells either passively (via cell death) or actively from living cells and is a well-established biomarker of cellular stress or injury (Trumpff et al., 2021). Under normal physiological conditions, apoptosis does not result in the release of free mtDNA into the cytosol. However, type III mitophagy (micromitophagy) in which mitochondria rid themselves of damaged components via vesicular release into the cytosol, is hypothesised to lead to increased ccf-mtDNA following both physical and psychological trauma (Thurairajah et al., 2018; Trumpff et al., 2019). However, at the whole population level, Blalock et al. (2024) reported no change in ccf-mtDNA levels when comparing combat trauma exposed male veterans with and without PTSD, perhaps indicating that the increase in ccf-mtDNA that occurs following trauma may only be present in the acute phase, directly following the inciting stressor. However, when controlling for age, diabetes status and the use of antidepressant medication, the PTSD group *was* observed to have lower ccf-mtDNA than the non-PTSD group (Blalock et al., 2024). Decreased levels of ccf-mtDNA have been suggested to reflect mitochondrial loss in the early stages of neurodegenerative diseases such as Parkinson’s and Alzheimer’s (Lowes et al., 2020; Podlesniy et al., 2013). Taken together, these findings suggest that exposure to trauma may be associated with mitochondrial dysfunction and increased ccf-mtDNA due to increased ROS production and cellular injury, but that chronic PTSD is defined by mitochondrial loss and thus a delayed decrease in ccf-mtDNA to a new, lower baseline level. Whilst further study is required to test this hypothesis,, it nevertheless highlights the importance of controlling for illness duration and time since the traumatic event(s) when studying the molecular consequences of PTSD.

The evidence strongly suggests the involvement of mitochondrial dysfunction and metabolic alterations in PTSD. These changes include impaired energy production, disrupted oxidative stress responses, and altered mitochondrial signaling, all of which contribute to neuroinflammation, neuronal damage, and the dysregulation of stress response pathways to exacerbate the chronic symptoms of PTSD (**Figure 2**).

**Fig. 2.**
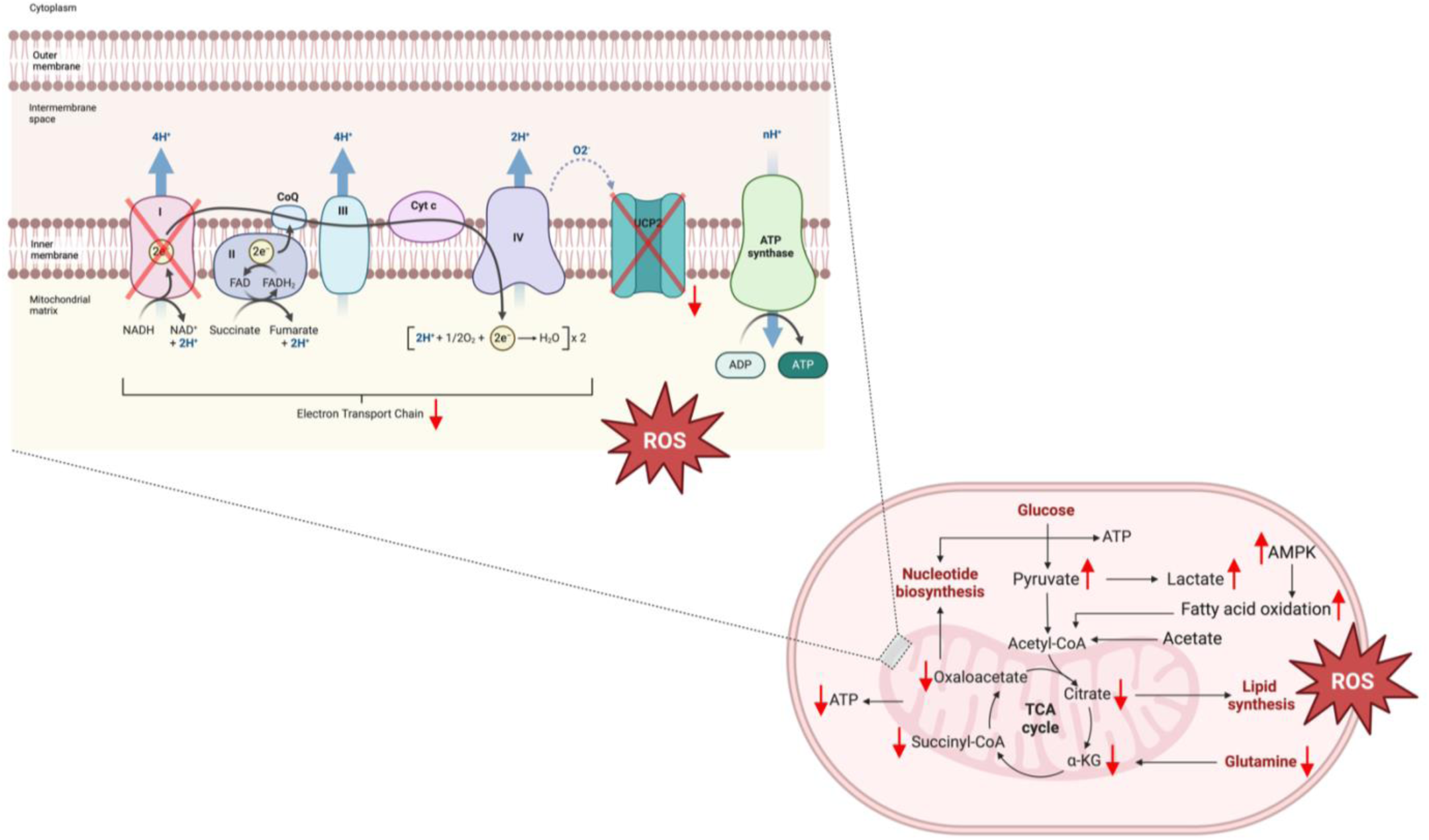
Suggested Mitochondrial Alterations in PTSD. Increased energy demand following trauma results in increased fatty acid oxidation to provide energy, but a corresponding increase in ROS which damage mitochondria. Downregulation of UCP2 further increases ROS production via effects on the mitochondrial membrane potential and decreases oxaloacetate levels. These alterations may be exacerbated by mtDNA mutations which alter genes encoding complex I of the electron transport chain, thus reducing ATP production. *Created in BioRender. Rye, C. (2025)* https://BioRender.com/v17a176.

### Treatments

Given the clear connection between mitochondrial and metabolic alterations and PTSD symptoms, addressing oxidative stress and mitochondrial health may offer promising therapeutic avenues for individuals with PTSD. We excluded traditional medicines such as Anshen Dingzhi prescription and hominis placenta and psychedelic compounds due to their broad and often less specific mechanisms of action, our search identified 12 preclinical studies exploring this possibility (Table 3). These could be divided into treatments targeting mitochondrial functioning via six mechanisms: i) energy production; ii) ROS production; iii) mitochondrial biogenesis; iv) apoptosis; v) inflammation, and; vi) neurosteroidogenesis. However, many appeared to target multiple, interrelated processes to ameliorate mitochondrial alterations. For example, in their investigation of risperidone treatment following SRS-exposure, Garabadu and colleagues (2015) reported that risperidone administered orally at 0.1mg/kg not only ameliorated PTSD-like behavioural symptoms and cognitive deficits, but also reversed the increased activity of the mitochondrial respiratory chain. Risperidone decreases activity of mitochondrial complex I (Balijepalli et al., 2001), and this is the likely mechanism for the risperidone-induced stabilisation of mitochondrial functioning in SRS-exposed animals. Furthermore, risperidone had anti-apoptotic effects, which are likely to increase cell viability and ATP production (Tsialtas et al., 2021). Since some anti-apoptotic mechanisms also have anti-inflammatory effects (Talanian et al., 2000), risperidone might further help to reduce the inflammation that is often seen in PTSD and thus prevent the vicious cycle of inflammation-induced mitochondrial dysfunction.

**Table 3:** Potential treatments for mitochondrial dysfunction in PTSD. *Abbreviations:* SPS = single prolonged stress; SPS+S = single prolonged stress + shock; SRS = stress, re-stress, MMP = mitochondrial membrane potential; ROS = reactive oxygen species; ETC = electron transport chain; TSPO = translocator protein; NLRP3 = nucleotide-binding domain and leucine-rich repeat protein-3.

| Author | Year | Species | Model | Treatment | Reported Mechanisms |
| --- | --- | --- | --- | --- | --- |
| Bhattacharjee et al. | 2021 | Rats | SRS | Taurine, Paroxetine | Cellular energy production, MMP |
| Chen et al. | 2024 | Mice | SPS | G1 | Cellular energy production, apoptosis, ROS |
| Garabadu et al. | 2015 | Rats | SRS | Risperidone | Apoptosis, ETC, MMP |
| Gong et al. | 2023 | Mice | SRS | Leptin | NLRP3 Inflammasome |
| Ji et al. | 2023 | Mice | SRS | Leptin | ROS, NLRP3 Inflammasome |
| Kao et al. | 2016 | Mice | Unsignalled footshock | Fluoxetine | Citric acid cycle |
| Mei et al. | 2023 | Rats | SPS | Sodium aescinate | NLRP3 Inflammasome, ROS, cellular energy production |
| Prajapati et al. | 2024 | Rats | SRS | Suvorexant |  |
| Wang et al. | 2017 | Rats | SPS | Metformin | ROS, mitochondrial biogenesis |
| Wang et al. | 2024 | Mice | SPS | Resveratrol | Cellular energy production, MMP |
| Xie et al. | 2024 | Mice | SPS+S | Cannabidiol | ETC |
| Zhang et al. | 2016 | Rats | Time-dependent sensitisation | AC-5216 | TSPO, neurosteroidogenesis |

Targeting cellular energy production and apoptotic pathways is not the sole mechanism by which to reverse the mitochondrial alterations observed in PTSD. Given the connection between oxidative stress, lipid peroxidation, and mitochondrial dysfunction in PTSD, strategies to reduce oxidative damage (Chen et al., 2024; Gong et al., 2023; Ji et al., 2023; Mei et al., 2023; Wang et al., 2017) also appear beneficial in treating or preventing PTSD. Indeed, via inhibiting activation of the NLRP3 inflammasome in astrocytes, sodium aescinate (SA) (Mei et al., 2023) and leptin (Gong et al., 2023; Ji et al., 2023) both appear to decrease mitochondrial ROS to protect against PTSD-like symptoms. Interestingly, however, they do so via distinct signalling pathways. Leptin effects are mediated by the Janus kinase/signal transducer and activator of transcription 3 (JAK2/STAT3) signalling cascade (Ji et al., 2023), while SA exerts an indirect protective effect on neurons by inhibiting peripheral inflammation (Ding et al., 2021), reducing levels of MDA and scavenging ROS (Huang et al., 2020). Metformin is also believed to target ROS production to protect against PTSD-like symptomatology; upregulating the expression of oxidative stress-related genes in the hippocampus and preventing SPS induced down-regulation of AMPK signalling (Wang et al., 2017). However, to our knowledge, metformin has only been studied as a prophylactic treatment in animal models, given before trauma exposure (Wang et al., 2017). This raises several clinical issues, primarily because there is currently no suitable biomarker by which to identify individuals at risk of developing PTSD. Moreover, even if prospective identification was possible, this would raise several ethical issues including the risk of stigmatising vulnerable populations or administering medication to individuals who may never develop PTSD. Additionally, the long-term effects of pre-emptive treatment with metformin in healthy individuals remain unclear, further complicating its use in this context.

An alternative approach currently under investigation is the targeting of neurosteroidogenesis (Zhang et al., 2016). This is largely due to the fact that the intracellular cholesterol reserve of steroidogenic cells controls the production of cortisol hormones, suggesting that the alterations in cortisol concentration that occur in PTSD are connected to steroidogenesis inside mitochondria (Dmytriv et al., 2023). Dysregulation of allopregnanolone synthesis - a neurosteroid with anxiolytic and neuroprotective properties (Kita et al., 2004) - has been implicated in heightened stress responding in PTSD (Rasmusson et al., 2006). One drug currently being investigated is AC-5216, a selective modulator of the GABA-A receptor (Zhang et al., 2016). AC-5216 acts to enhance GABA-A receptor activity to indirectly stimulate the production of neurosteroids, reduce HPA axis hyperactivity and alleviate neuroinflammation (Kita et al., 2004). Furthermore, via effects on mitochondrial efficiency, AC-5216 may mitigate oxidative stress and support cellular repair mechanisms (Skokou et al., 2023), helping to reverse some of the neurobiological changes observed in PTSD. This targeted action on both mitochondrial health and neurosteroidogenesis offers a dual approach to combating the underlying pathophysiological changes in PTSD, potentially leading to improved symptom management and recovery.

Clearly there are several promising avenues via which to develop treatments for PTSD which specifically target mitochondrial dysfunction. However, it must be noted that many of these are only just starting to be investigated in the preclinical space, meaning they may not be available for several years. Compounding this is the fact that psychiatry has the second-lowest success rate of drugs entering the marketplace, with a 94% chance of failure after entering Phase I trials (Mullard, 2016). Consequently, existing medications such as risperidone, which are already approved for psychiatric conditions like schizophrenia, may offer a more immediate translational pathway. These drugs, with established safety profiles and regulatory approval, could, following appropriate preclinical and clinical trials to demonstrate efficacy, potentially be repurposed for PTSD treatment, facilitating their quicker integration into clinical practice.

## CONCLUSION

Data from both pre-clinical and clinical studies strongly implicates mitochondrial and metabolic processes in the complex pathogenesis of PTSD, suggesting that PTSD may be better conceptualised as a systemic illness with substantial somatic manifestations. Here we have reviewed several articles exploring mitochondrial alterations following trauma. While the majority of the studies show a link between PTSD and oxidative stress leading to apoptosis, contradictory results concerning the nature of alterations to the mitochondrial respiratory chain suggest that the mechanisms behind this association are still not fully understood. Indeed, it remains contested as to whether these mitochondrial and metabolic pathologies are a risk factor for the development of PTSD or result from PTSD itself. Further research to distinguish these possibilities will be required if we are to identify and develop novel treatments which specifically target mitochondrial dysfunction in PTSD – either to improve PTSD symptoms or to treat comorbid somatic conditions.

## Data Availability

The data used in this review are publicly available and specific references to the datasets and data sources used are provided in the references section of the manuscript.

https://osf.io/9az8p

## Acknowledgements

FEA is supported by UK Research and Innovation’s Biological and Biotechnology Research Council grant BB/W001195/1. CSR is jointly supported by a Machin studentship and Q550 studentship from Queens’ College, University of Cambridge. ALM is the Ferreras-Willetts Fellow in Neuroscience at Downing College, University of Cambridge. For the purpose of open access, the authors have applied a Creative Commons Attribution (CCBY) licence to any Author Accepted Manuscript version arising from this submission.

## Author Contributions

**Felippe E. Amorim:** Conceptualisation, Methodology, Investigation, Visualisation, Formal Analysis, Writing – Original Draft, Review & Editing.

**Charlotte S. Rye:** Conceptualisation, Data Curation, Methodology, Investigation, Visualisation, Formal Analysis, Writing – Original Draft, Review & Editing.

**Amy L. Milton:** Funding acquisition, Supervision, Writing – Review & Editing.

## Statements and Declarations

### Ethical Considerations

All included preclinical studies were approved by the competent authority responsible for ensuring compliance with the regulations governing the use of animals in scientific experiments and awarded ethical approval by the official body of the respective country. All included clinical studies were approved by ethical committees at the respective institutions, and protocols complied with national legislation, with the privacy rights of all participants observed.

### Consent to Participate

NA

### Consent for Publication

All authors have reviewed and approved the manuscript and agree to its submission for publication in the Journal of Psychopharmacology.

### Declaration of Conflicting Interests

The authors declare no conflicting interests.

### Funding Statement

This systematic review was supported by the UK Research and Innovation’s Biological and Biotechnology Research Council under grant number BB/W001195/1. Although no direct financial costs were incurred during the course of this review, the grant provided administrative and research support.

## Notes

### Competing Interest Statement

The authors have declared no competing interest.

### Funding Statement

This systematic review was supported by the UK Research and Innovations Biological and Biotechnology Research Council under grant number BB/W001195/1. Although no direct financial costs were incurred during the course of this review the grant provided administrative and research support.

### Author Declarations

The study used ONLY openly available human data from primary research articles available on PubMed, Web of Science and Scopus.

